# Effects of repeated vaccination and vaccine formulation on the induction of broadly neutralizing antibody responses against influenza A virus in children

**DOI:** 10.1101/2021.08.03.21261495

**Authors:** Sergey Yegorov, Daniel B. Celeste, Kimberly Braz Gomes, Jann C. Ang, Colin Vandenhof, Joanne Wang, Ksenia Rybkina, Vanessa Tsui, Mark Loeb, Matthew S. Miller

## Abstract

The induction of broadly neutralizing antibodies (bNAbs) that target the hemagglutinin stalk domain is a promising strategy for the development of “universal” influenza virus vaccines. bNAbs can be boosted in adults by sequential exposure to heterosubtypic viruses through natural infection or vaccination. However, little is known about if/how bNAbs are induced by vaccination in more immunologically naive children. Here, we describe the impact of repeated seasonal influenza vaccination and vaccine type on induction of bNAbs in a well-characterized paediatric cohort enrolled in a series of randomized control trials of seasonal influenza vaccination. Repeated seasonal vaccination resulted in significant boosting of a durable bNAb response. Boosting of serological bNAbs titers was comparable within inactivated and live attenuated (LAIV) vaccinees and declined with age. These data provide new insights into vaccine-elicited bNAb induction in children, which has important implications for the design of “universal” influenza vaccine modalities in this critical population.

## INTRODUCTION

Despite decades of study and the accessibility of seasonal vaccination, influenza remains an important public health concern, largely due to its ability to escape immunity and cause seasonal epidemics and pandemics ^1–3^. Seasonal vaccines, the mainstay of influenza prevention, have often shown modest real-world effectiveness due to suboptimal immunogenicity, alongside rapid and unpredictable changes in circulating virus strains ^1–3^. Globally, several different seasonal influenza vaccine formulations are approved, including intramuscular injection of inactivated virus (IIV) or mucosal administration of replication-competent live-attenuated virus (LAIV) ^1^. Although influenza vaccines are often considered equally efficacious, there are insufficient data on the impact of repeated vaccination and vaccine type on the breadth of influenza virus-directed immunity in specific demographic groups, especially children, who are a major source of influenza transmission and alongside the elderly, are at heightened risk for influenza-related hospitalizations and death ^4,5^.

The bulk of influenza vaccine-elicited antibodies (Abs) target the viral membrane surface proteins haemagglutinin (HA), and to a lesser extent, neuraminidase (NA). Of the HA-reactive Abs, most bind the “immunodominant” globular HA head domain in a strain-specific manner, blocking the interaction between the virus and cell surface sialic acid, while a minority of HA-reactive Abs bind the sub-dominant, but highly conserved HA stalk domain; a subset of these antibodies is considered broadly-neutralizing (bNAbs). HA stalk-reactive bNAbs have emerged as a promising strategy for the development of a universal influenza vaccine ^6,7^.

In adults, we and others have shown that bNAbs are selectively boosted by exposure to pandemic/heterosubtypic HA subtypes (via either infection or vaccination)^8–10^. bNAb titers also increase over time due to an accumulation of lifetime exposures to influenza virus selecting for cross-reactive B cell clone maintenance and expansion^11,12^. A recent clinical trial demonstrated that chimeric hemagglutinin vaccines delivered in live-attenuated and inactivated formulations could successfully boost bNAb titers in adults ^6,7^. However, little is known about how bNAbs develop in children, and how vaccination impacts bNAb development-knowledge that will have important implications for understanding the nature of vaccine-mediated immunity against influenza, especially in the context of exposure to novel strains ^1,13,14^.

Therefore, we explored the effects of seasonal influenza vaccination and vaccine formulation on bNAb induction in participants of a cluster-randomized control trial (cRCT) designed to compare the community-level protection mediated by IIV and LAIV vaccination of children. We found that titers of neutralizing bNAbs were enhanced by repeated seasonal vaccination, and that bNAb levels were boosted similarly by LAIV and IIV. As expected, LAIV and IIV effects on bNAb titers were more pronounced in the mucosa and blood, respectively, and the magnitude of vaccine-induced bNAb responses in the serum declined with age.

## RESULTS

### Study participants

The effects of repeated seasonal IIV vaccination on the induction of bNAbs were studied using serum samples from 68 RCT participants (median age 9.0 years, Table 1), who received either IIV (37 “vaccinees”), or 31 controls, for each of the three seasons (2008/09 to 2010/11) captured by the cRCT (Fig 1A and Table 3), and for whom paired samples were available. The effects of vaccine type on bNAb elicitation were subsequently studied using serum and mucosal samples from 72 RCT participants (median age 11.0 years, Table 2), who received either the inactivated vaccine (“IIV”, n=35), or the live attenuated vaccine (“LAIV”, n=37) (Fig 1B and Table 3), and for whom paired samples were available.

**Table 1:** Demographic characteristics of study participants in analysis I (Effect of repeated seasonal influenza vaccination on bNAb titers in children).

|  | All (68) | Controls (31) | IIV3 vaccinees (37) | P value |
| --- | --- | --- | --- | --- |
| Median age, years [IQR] | 9.0 [6.0;11.0] | 8.0 [5.0;11.0] | 9.0 [6.5;11.0] | 0.484 |
| Male sex, n (%) | 38 (55.9%) | 19 (61.3%) | 19 (51.4%) | 0.676 |

IQR=interquartile range
Age and sex differences were assessed using the two-sided Mann-Whitney U and Fisher's exact tests, respectively.

**Table 2:** Demographic characteristics of study participants in analysis II (Effect of vaccine type (IIV vs. LAIV) on induction of bNAbs in children).

|  | All (68) | IIV3 vaccinees (35) | LAIV3 vaccinees (37) | P value |
| --- | --- | --- | --- | --- |
| Median age, years [IQR] | 11.0 [8.0;13.0] | 11.0 [8.0;13.0] | 11.0 [9.0;13.0] | 0.768 |
| Male sex, n (%) | 36 (52.9%) | 19 (54.2%) | 17 (45.9%) | 0.638 |

IQR=interquartile range
Age and sex differences were assessed using the two-sided Mann-Whitney U and Fisher's exact tests, respectively.

**Table 3:** Description of influenza strains used in the standard-of-care trivalent vaccines administered over the study period.

|  | Season | Strain Type | Strain Subtype | Strain name |
| --- | --- | --- | --- | --- |
| *Analysis I*<br>*Seasons* | 2008-09 | A | H1N1 | Brisbane/59/2007 |
|  |  |  | H3N2 | Brisbane/10/2007 |
|  |  | B | Yamagata | Florida/4/2006 |
|  | 2009-10 | A | H1N1 | Brisbane/59/2007 |
|  |  |  | H3N2 | Brisbane/10/2007 |
|  |  | B | Victoria | Brisbane/60/2008 |
|  | 2010-11 | A | H1N1 | California/7/2009 |
|  |  |  | H3N2 | Perth/16/2009 |
|  |  | B | Victoria | Brisbane/60/2008 |
| *Analysis II*<br>*Seasons* | 2014-15 | A | H1N1 (pdm09) | California/7/2009 |
|  |  |  | H3N2 | Texas/50/2012 |
|  |  | B | Yamagata | Massachusetts/2/2012 |

**Fig 1.**
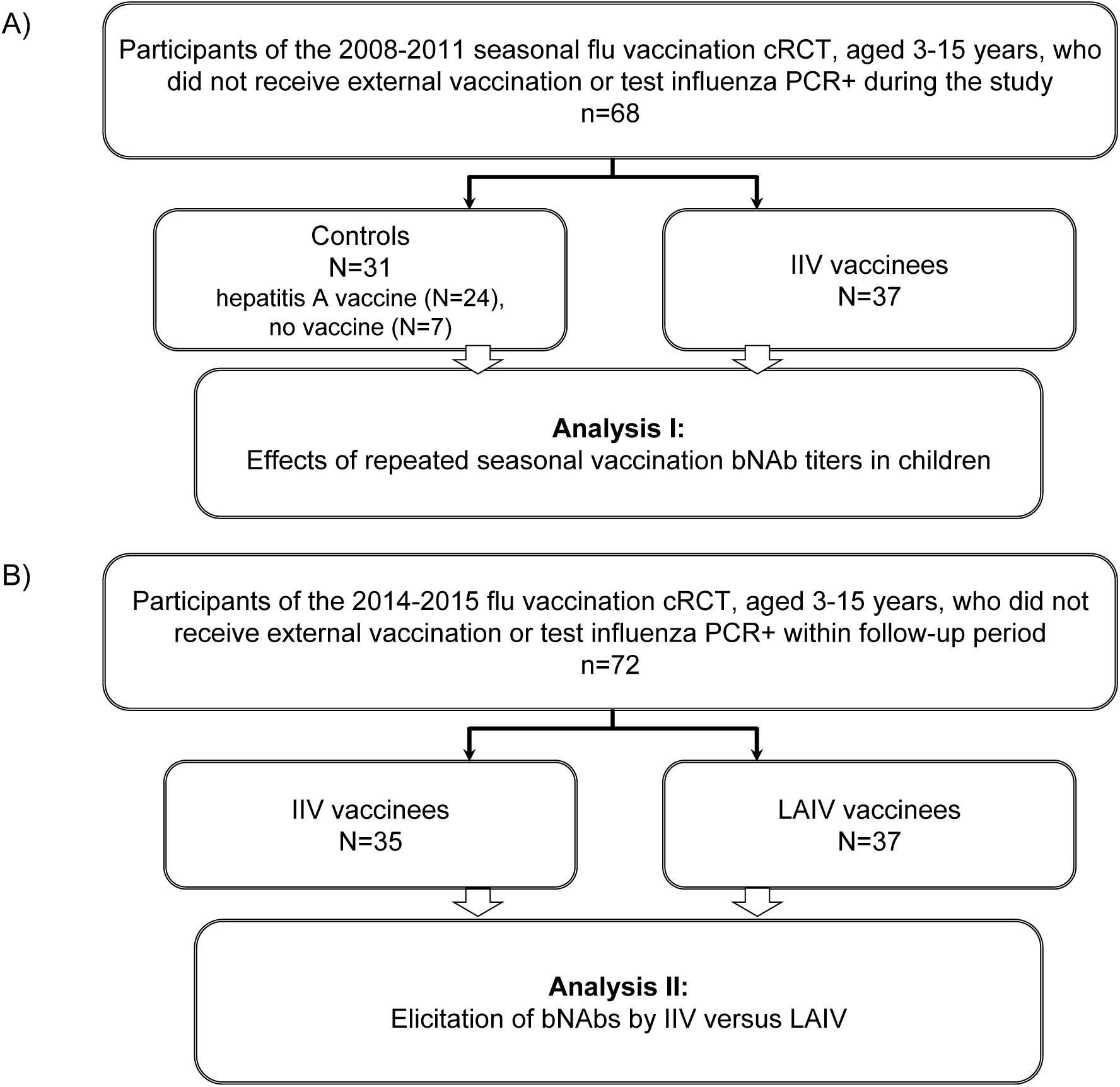
Study flow chart. A) Description of participants in analysis I: “Effects of repeated seasonal influenza vaccination on bNAb titers in children “. B) Description of participants in analysis II: “Effect of vaccine type (IIV vs. LAIV) on induction of bNAbs in children”. cRCT: cluster randomized controlled trial; bNAb: broadly neutralizing antibodies; IIV: inactivated influenza vaccine; LAIV: live attenuated influenza vaccine.

### Effects of repeated seasonal vaccination on serologic bNAb responses

To determine the effect of repeated seasonal IIV vaccination of children on the induction of bNAbs, microneutralizaton (MNT) assays were performed on the 2008/09 and 2012/13 pre-vaccination sera (corresponding to the beginning of the cRCT, and one year after its conclusion, respectively) using cH5/1 N3, a chimeric virus previously validated to quantify neutralization mediated by group 1 HA stalk-binding bNAbs ^9,10,13,14^. After three vaccination seasons, higher MNT_50_ titers were evident in vaccinees compared to controls (geometric mean fold change (GMFC) = 2.65 vs. 1.43, p < 0.001, Fig 2A, C); 43 % (16/37) of vaccinees had at least a fourfold increase of MNT_50_, compared with the 13 % (4/31) seroconversion among controls. Hemagglutination inhibition (HAI) titers against Cal/09, which was the H1N1 component of the seasonal influenza vaccine beginning in 2010/11^15^ (see Table 3) were slightly elevated after repeated seasonal vaccinations (19/37, 51 % had at least a fourfold increase of HAI), but the post-vaccination HAI titers in most (29/37) vaccinees remained below 40 (the conventional seroprotection threshold), and the change was not significantly different between vaccinees and controls (GMFC=5.18 vs. 3.64, p=0.308, Fig 2B,D). The magnitudes of vaccine-elicited MNT_50_ or HAI boosting were not correlated with age (Fig E-F). These results demonstrate that repeated seasonal IIV vaccination induces durable bNAb responses in children.

**Fig 2.**
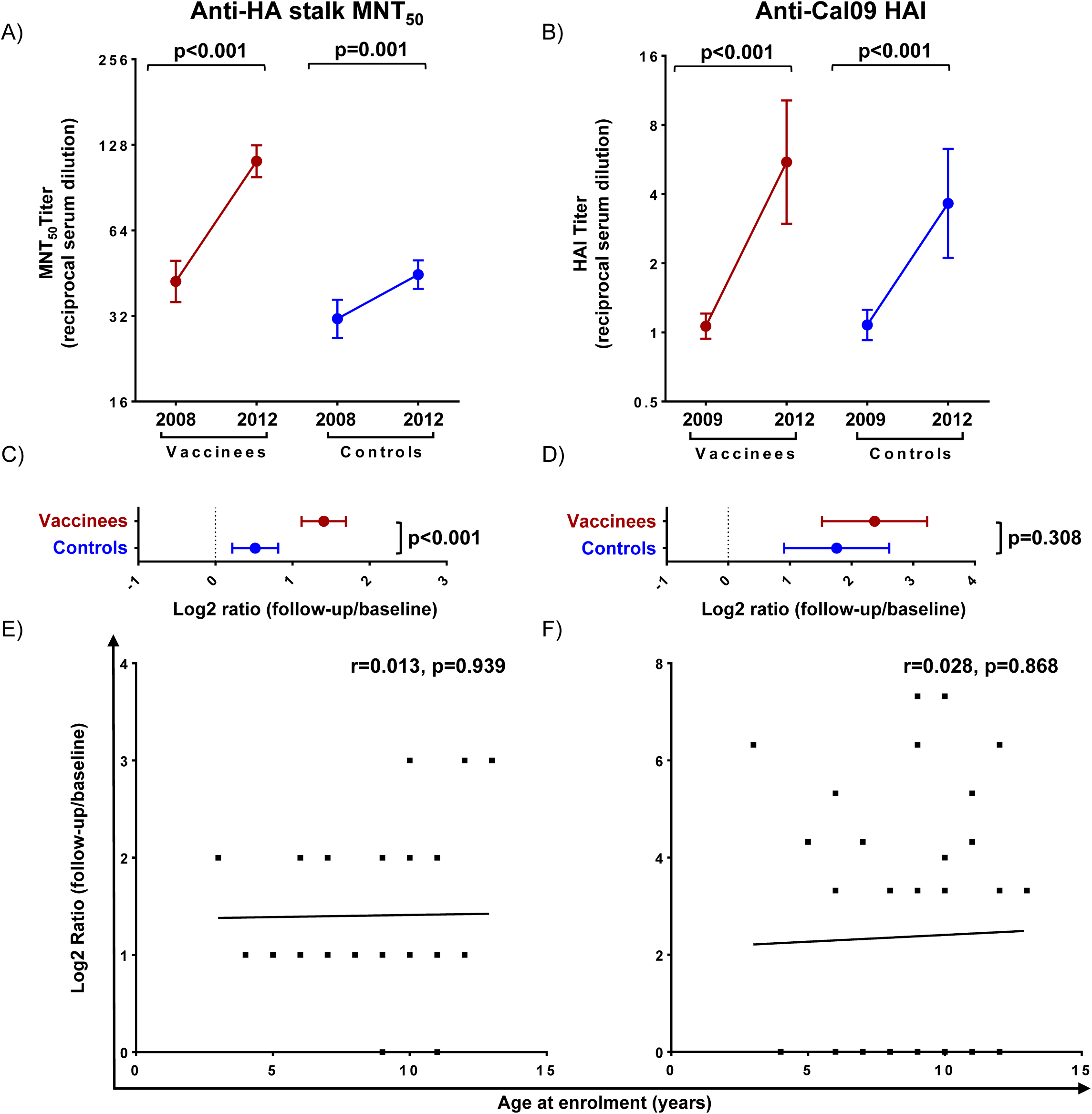
Effects of seasonal vaccination of bNAb titers in children. A) Anti-HA stalk MNT_50_ titers at baseline (year 2008) and after 3 cRCT vaccination seasons (year 2012). B) Serum HAI activity against the Cal/09 virus assessed at baseline (year 2009) and after 3 cRCT vaccination seasons (year 2012). C) Log2-transformed ratios of MNT_50_ titers post-repeated vaccination (year 2012) versus at baseline (year 2008). D) Log2-transformed ratios of HAI titers post-repeated vaccination (year 2012) versus at baseline (year 2009). E-F) Correlation plots of log2-transformed MNT_50_ (E) and HAI (F) titer ratios and participant ages at enrollment. In panels A-B: dots and brackets represent geometric means and 95% confidence intervals, respectively; P values indicate the statistical significance of the intra-individual difference between pre- and post-vaccination titers assessed by paired t-test. In panels C-D: dots represent means, brackets depict 95% confidence intervals; P values indicate the statistical significance of the difference between vaccinees and controls assessed by unpaired t-test. In panels E-F: the Pearson coefficients (r) and their statistical significance are shown along with lines of best fit for vaccinees.

### Effects of vaccine type on serological bNAb responses

IIV and LAIV vaccines are known to elicit distinct immune responses ^16–18^. Thus, we set out to determine whether vaccine formulation (IIV vs. LAIV) influences the magnitude of serological vaccine-induced bNAb responses in children. There was a significant elevation of HA stalk-binding (anti-cH6/1) IgG and IgA, and bNAb (anti-cH5/1 N3) MNT_50_ titers at one-month post-vaccination (GMFC=2.71, p<0.001; 1.51, p=0.004; 1.48, p=0.002, respectively, Fig 3A-C); this elevation was more pronounced in the IIV vaccinees than in LAIV vaccinees, although the response magnitude was not significantly different between the two groups (Fig 3D-F). The magnitude of vaccine-elicited boosting of both anti-HA stalk IgG and MNT_50_ (r=-0.238, p=0.045 and r=-0.333, p=0.004, respectively), but not anti-HA stalk IgA titers, declined with age (Fig 3G-I). Altogether, these data show that in young children, both IIV and LAIV boost serum bNAbs in an age-dependent manner.

**Fig 3.**
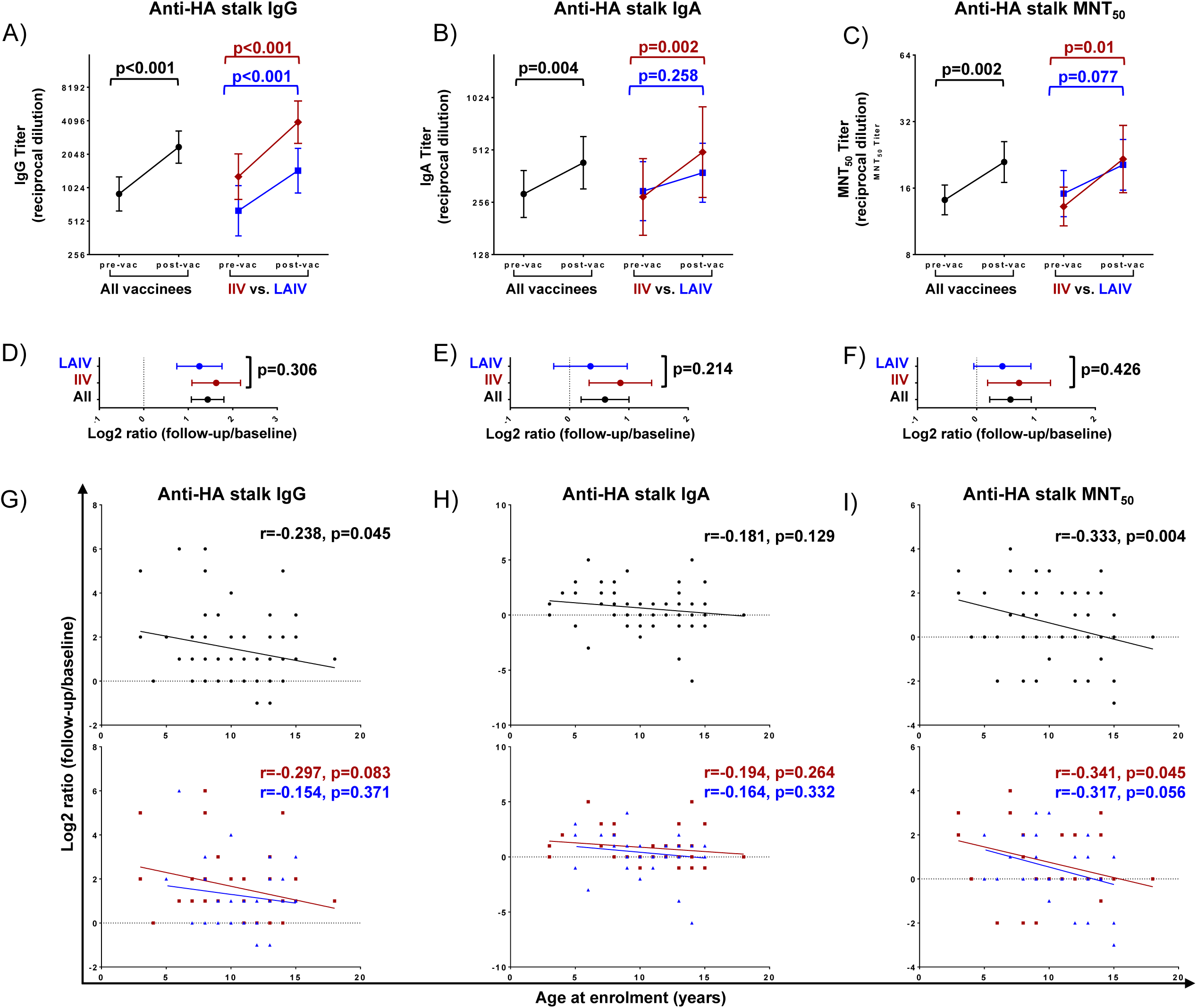
Effect of vaccine type on serological bNAb titers. A-C) Anti-HA stalk IgG (A), IgA (B) and MNT_50_ (C) titers pre- and post-vaccination. D-F) Log2-transformed ratios of anti-HA stalk IgG (D), IgA (E) and MNT_50_ (F) titers post- versus pre-vaccination. G-I) Correlation plots of log2-transformed anti-HA stalk IgA (G), anti-HA stalk IgG (H) and MNT_50_ (I) titer ratios and participant ages at enrolment. In panels A-C: dots and brackets represent geometric means and 95% confidence intervals; P values indicate the statistical significance of the intra-individual difference between pre- and post-vaccination titers assessed by paired t-test. In panels D-F: dots and brackets represent means and 95% confidence intervals, respectively; P values indicate the statistical significance of the difference in the magnitude of immune response between IIV and LAIV vaccinees assessed by unpaired t-test. In panels G-I: the Pearson coefficients (r) and their statistical significance are shown along with lines of best fit for all participants (black) and IIV (red) versus LAIV (blue) vaccinees.

### Effects of vaccine type on strain-specific serological responses

We next asked whether seasonal influenza vaccine type influenced the magnitude of serological responses against strain-specific antigens included in the vaccine. Both IIV and LAIV induced a significant increase of anti-Cal/09 H1N1 IgG, IgA, and HAI titers at one-month post-vaccination (GMFC=3.0, 1.48 and 3.04, respectively, p<0.001 for all, Fig 4A-C). The boost in anti-Cal09 H1N1 IgG titers was significantly higher in the IIV group compared to the LAIV vaccinees (GMFC=4.59 vs. 2.0, p=0.017, Fig 4D). There was no difference in magnitude of boosting between the two groups for Cal-09 H1N1 IgA or HAI responses, though the absolute post-boost HAI titers were higher for IIV vaccinees (Fig 4E-F). The magnitude of vaccine-elicited Cal/09 H1N1 serum IgA increase was inversely correlated with age in all vaccinees (r=-0.379, p=0.001, Fig 4H). Boosting of Cal/09 H1N1 HAI was also inversely correlated with age for IIV vaccinees (r=-0.409, p=0.015, Fig 4I). Thus, the boost in functional antibody responses against vaccine antigen appears more pronounced in IIV and exhibits some age-dependence.

**Fig 4.**
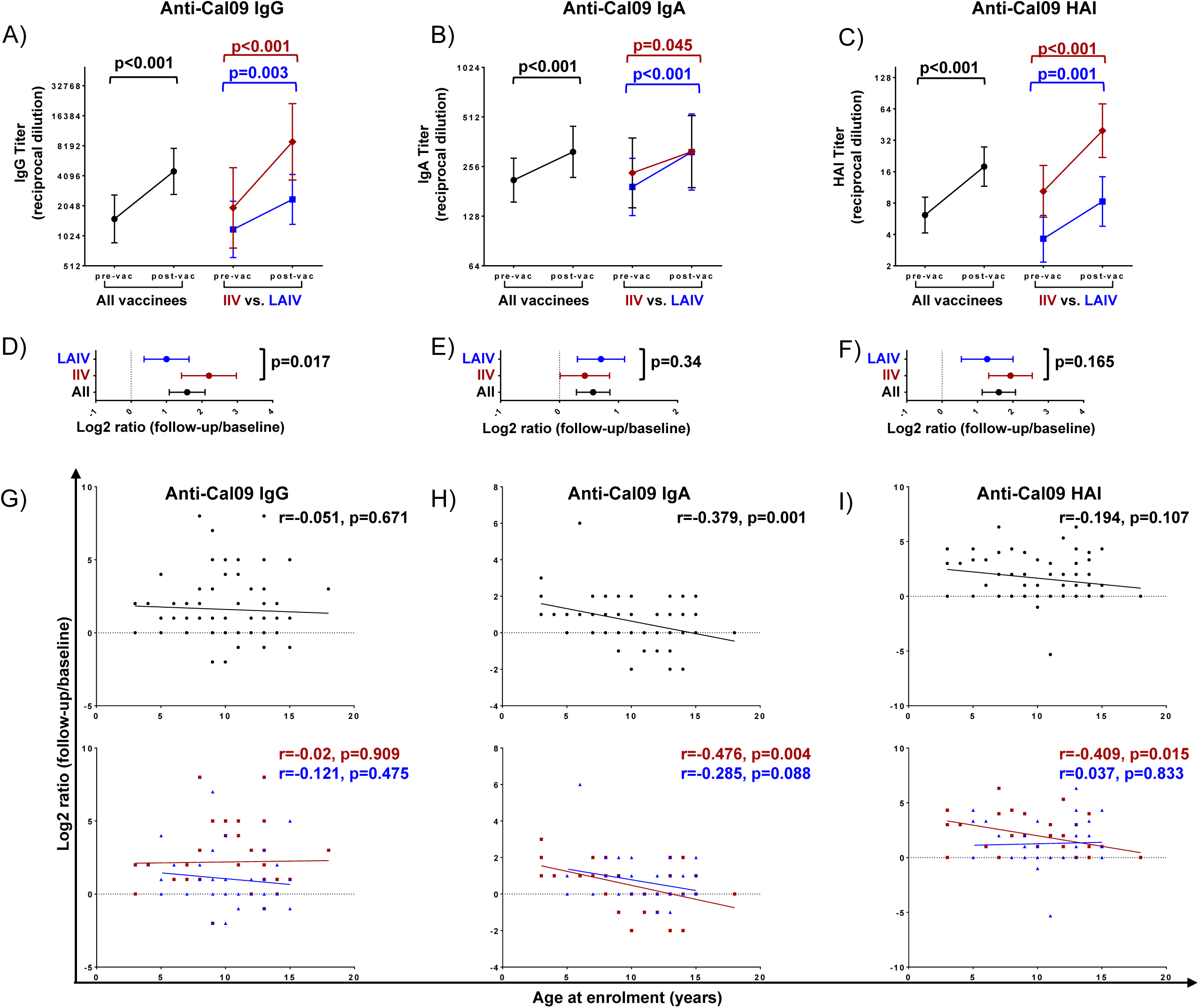
Effect of vaccine type on serological titers of strain-specific antibodies. A-C) Anti-Cal/09 IgG (A), IgA (B) and HAI (C) titers pre- and post-vaccination. D-F) Log2-transformed ratios of anti-Cal09 IgG (D), IgA (E) and HAI (F) titers post- versus pre-vaccination. G-I) Correlation plots of log2-transformed anti-Cal09 IgA (G), anti-Cal09 IgG (H) and HAI (I) titer ratios and participant ages. In panels A-C: dots and brackets represent geometric means and 95% confidence intervals; P values indicate the statistical significance of the intra-individual difference between pre- and post-vaccination titers assessed by paired t-test. In panels D-F: dots and brackets represent means and 95% confidence intervals, respectively; p values indicate the statistical significance of the difference in the magnitude of immune response between IIV and LAIV vaccinees assessed by unpaired t-test. In panels G-I: the Pearson coefficients (r) and their statistical significance are shown along with lines of best fit for all participants (black) and IIV (red) versus LAIV (blue) vaccinees.

### Effects of vaccine type on mucosal IgG and IgA responses

Vaccine-elicited mucosal immune responses are critical for protection against influenza ^1^. Therefore, we next assessed the antibody profiles in nasal swabs to determine the impact of vaccine type on mucosal immunity. Overall, vaccination tended to increase mucosal bNAb titers, without significant differences between vaccine types (Fig 5A-B, E-F). Anti-stalk IgG was significantly elevated, with the largest effect size in LAIV vaccinees (GMFC=3.17, p<0.001, Fig 5A, E). Notably, alongside an apparent increase in IgG there was a decrease in IgA titers against the Cal/09 H1N1 vaccine antigen in LAIV vaccinees, as reported previously ^16^. No significant correlations were observed between vaccine-mediated boosting of mucosal antibodies and age (Fig 5I-M). Altogether, LAIV and IIV boosted mucosal IgG and IgA bNAbs to a similar extent.

**Fig 5.**
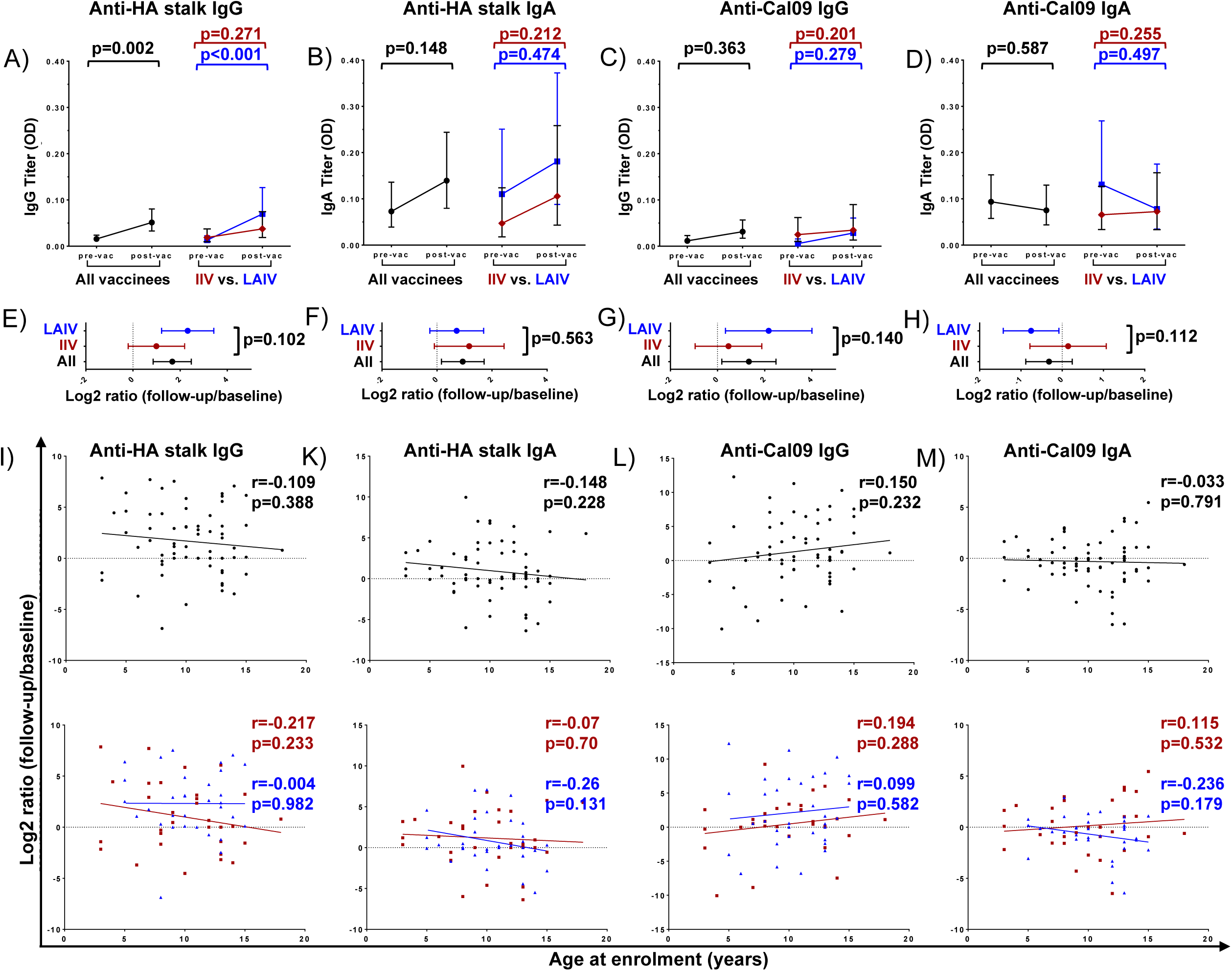
Effect of vaccine type on mucosal bNAb titers. A-D) Anti-HA stalk IgG (A), IgA (B), and anti-Cal/09 IgG (C) and IgA (D) titers. E-H) Log2-transformed ratios of anti-HA stalk IgG (E) and IgA (F), and anti-Cal09 IgG (G) and IgA (H) titers post- versus pre-vaccination. I-L) Correlation plots of anti- H6/1 IgG (E) and IgA (F), and anti-Cal09 IgG (G) and IgA (H) titer ratios and participant ages at enrolment. In panels A-D: dots and brackets represent geometric means and 95% confidence intervals; P values indicate the statistical significance of the intra-individual difference between pre- and post-vaccination titers assessed by paired t-test. In panels E-H: dots and brackets represent means and 95% confidence intervals, respectively; p values indicate the statistical significance of the difference in the magnitude of immune response between IIV and LAIV vaccinees assessed by unpaired t-test. In panels I-L: the Pearson coefficients (r) and their statistical significance are shown along with lines of best fit for all participants (black) and IIV (red) versus LAIV (blue) vaccinees.

### Relationship between vaccine-elicited serologic and mucosal antibody response

Whether and how vaccine-elicited immunity changes correlate between the systemic and mucosal compartments is a topic of debate ^19,20^. Therefore, we next explored the relationship between the magnitude of vaccine-induced bNAb boost in the serum versus the mucosa. In unstratified analysis, no correlation was observed between the serological and mucosal antibody responses (Fig 6A-D, left panels). Upon stratification by vaccine type, there was no correlation between serum and mucosal responses for bNAbs (Fig 6A-B, right panels). However, the magnitude of change in vaccine Cal/09 H1N1-reactive IgG, but not Cal/09 H1N1-reactive IgA, positively correlated between serum and mucosa in IIV vaccinees only (r=0.442, p=0.011, Fig 6C-D, right panels). Thus, bNAb responses appear to be compartment-specific.

**Fig 6.**
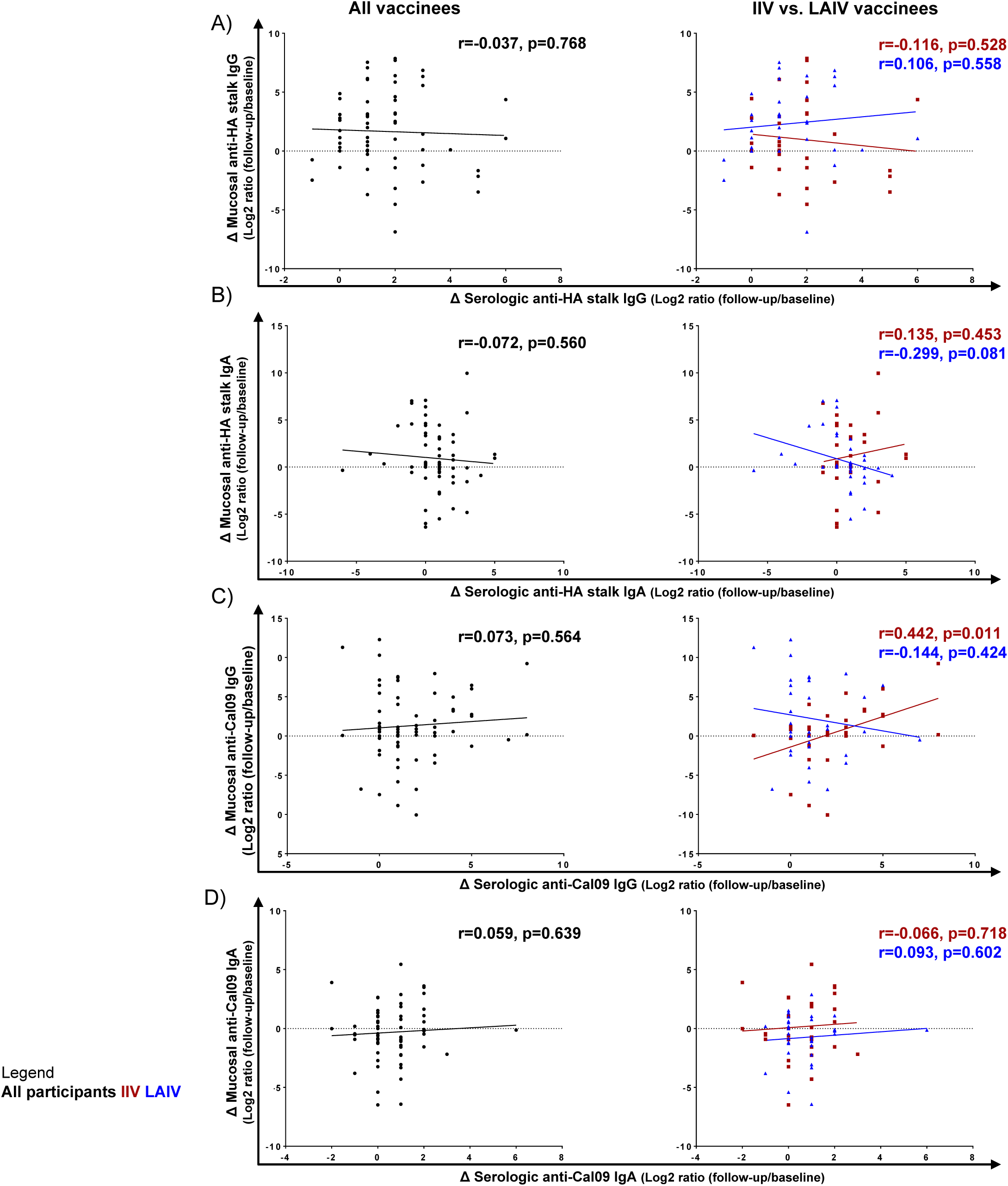
Relationship between mucosal and serological antibody titers. A-D) Correlation plots of anti- HA stalk IgG (A) and IgA (B), and anti-Cal09 IgG (C) and IgA (D) log2-transformed post-/pre- vaccination titer ratios. The Pearson coefficients (r) and their statistical significance are shown along with lines of best fit for all participants (black) and IIV (red) versus LAIV (blue) vaccinees.

## DISCUSSION

Notwithstanding the recent global decline in seasonal influenza incidence as a consequence of public health measures ^21^, influenza remains an important public health concern due to its inevitable seasonal re-emergence as social distancing eases, and the ever-present risk of future pandemics. Young children are a major driver of virus transmission, accounting for a significant proportion of influenza-related morbidity and mortality ^4,5^. Therefore, understanding how seasonal vaccination and varied vaccine formulations shape childhood immunity to influenza A viruses is of critical importance for effective influenza prevention. Vaccine-elicited immune responses are tightly linked to history of exposure to virus, infection and/or vaccination ^1,3^, and > 50 % of children have encountered influenza virus by 6 years of age ^22^. A major strength of our study is that it describes, for the first time to our knowledge, the impact of seasonal vaccination on bNAb induction in children whose exposure histories were carefully monitored within a cRCT framework, which allowed us to avoid confounding effects of infection and focus on the influence of vaccination, specifically.

After three vaccination seasons, bNAb-mediated virus neutralization in IIV vaccinees increased by > 2.5-fold, supporting the notion that seasonal vaccination facilitates sustained expansion of cross-reactive Ab repertoires ^10^. In contrast, although vaccine-elicited strain-specific HAI activity also rose over three seasons, it was not significantly different between the vaccinated and unvaccinated children at the conclusion of the study.

The magnitude of bNAb expansion induced by repeated IIV vaccination was inversely correlated with age, consistent with preferential boosting of antibodies against the HA head domain that is known to occur in adults after seasonal influenza vaccination ^9,23^. Notably, a very modest bNAb elevation was observed in control subjects between 2008 and 2012, which is consistent with the observation that HA stalk-reactive bNAbs tend to be polyreactive, and that polyreactive antibodies tend to increase with age ^24^. In contrast, HAI titer increases observed in vaccinees and controls during the study period were very similar, and overall, these titers remained very low – below the typical seroprotection threshold of 40. Together, these data suggest that seasonally repeated vaccination preferentially induced bNAbs in young children.

Importantly, IIV and LAIV were similarly effective at boosting bNAbs in children, in line with the equivalent efficacy offered by these two vaccine types in the cRCT ^25^. Furthermore, vaccine-elicited bNAb boosting was more pronounced in the blood of IIV vaccinees, and in the mucosa of LAIV vaccinees, consistent with the respective vaccination routes.

Mucosal immunity plays a critical role in protection against respiratory infections. The upper respiratory tract antibody repertoire is dominated by secretory IgA, while in the lower respiratory tract IgG is more abundant ^1^. We have previously shown that broadly-neutralizing IgA is more potent than IgG ^13^. This is, at least in part, due to the glycosylation of IgA, whereby sialic acids present on the antibody mediate a second mode of binding that inhibits viral entry ^26^. We have also recently shown the influenza virus-IgA immune complexes signal through the FcαRI receptor on neutrophils to stimulate NETosis, and that neutrophil extracellular traps are capable of inactivating influenza virus ^27,28^. Therefore, robust mucosal immune response stimulation should be a priority for the development or more broad and efficacious influenza vaccines.

Interestingly, we did not observe a correlation between bNAb responses in the blood and nasal mucosa, suggesting an important contribution from *in situ* antibody generation at the mucosa. Furthermore, while the serum bNAb and Cal/09-specific IgA titers were similar, the titers of Cal/09-reactive IgG were approximately two-fold higher compared to HA stalk-reactive IgG, consistent with the immunodominant nature of the HA head domain ^1^. Notably, mucosal titers of bNAbs and strain-specific Abs were similar, suggesting that mucosal responses may be less focused on immunodominant epitopes.

### Study Limitations

First, our analyses were based on a relatively small selection of samples derived from the original cRCTs due to sample availability. Second, for reasons of feasibility we assessed only bNAbs against group 1 HAs. In the future, it would be important to examine group 2 bNAbs as well. Our mucosal analyses were performed using nasal swabs which precluded conventional antibody titrations due to the low antibody levels typical for those samples. However, we have shown previously that we are able to reliably measure differences in mucosal IgA using the methods employed herein ^16^.

### Conclusion

Our data provide unique insights into the effects of repeated seasonal influenza vaccination and vaccine type on bNAb induction in children. Seasonal influenza vaccines in children are capable of inducing bNAbs against influenza A both in blood and respiratory mucosa, which has important ramifications for the selection of “universal” vaccine platforms that could be effectively deployed in children. Future studies will need to explore factors responsible for enhanced bNAb generation in the context of seasonal vaccination, how vaccine-elicited bNAbs functionally relate to those induced by natural infection ^29^, and whether bNAb induction mechanisms can be utilized to improve “real world” effectiveness of influenza vaccines.

## METHODS

### Study setting and participant recruitment

This study was nested within the multi-centre cluster randomized controlled trials (cRCT) of annual influenza vaccination in Hutterite communities ^25,30,31^. Selection of samples from the cRCT was done randomly, ensuring that the comparison groups were age- and sex-matched (see Tables 1 and 2). Specifically, we included participants with a complete history of repeated vaccinations, who did not receive any external influenza vaccines and never tested PCR-positive for influenza A or B within the study period. For analysis of repeated vaccination with IIV (Fig 1A), we used annually collected pre-vaccination samples from the 2008-09, 2009-10 and 2012-13 seasons. The control group for this study included individuals who either received a hepatitis A vaccine, or no vaccine. No differences in immune responses were detected between the hepatitis A-vaccinated participants and the unvaccinated participants, and so they were grouped together for the purposes of this study. To assess the impact of vaccine formulation (IIV vs. LAIV, Fig 1B) we used samples collected at pre-vaccination day 0 and post-vaccination day 28 during the 2014-15 season. Vaccines were administered annually in October; participants’ records were screened for a period of 3 months post-vaccination to allow for reporting of any external vaccination and for reporting of PCR-confirmed influenza. The IIV and LAIV vaccinees received a 0.5-mL intramuscular injection of Vaxigrip (Sanofi Pasteur) or a 0.2-mL dose of intranasal FluMist (MedImmune), respectively. The following influenza strains were used in the standard-of-care trivalent vaccines administered over the study period: A/Brisbane/59/2007(H1N1), A/Brisbane/10/2007 (H3N2), B/Florida/4/2006-like virus (2008-09); A/Brisbane/59/2007 (H1N1), A/Brisbane/10/2007 (H3N2) and B/Brisbane/60/2008 (2009-10); A/California/7/2009 (H1N1), A/Perth/16/2009 (H3N2), B/Brisbane/60/2008 (2010-11) and A/California/7/2009 (H1N1)pdm09, A/Texas/50/2012 (H3N2), B/Massachusetts/2/2012 (2014-15) ^25,30,31^ (Table 3). The control group consisted of participants who received either hepatitis A vaccine (N=24) or no vaccine (N=7) during the study period (Fig 1A); no differences in any of the assessed demographic or immune parameters were seen between these two subsets within the control group. All study procedures were approved by the McMaster University Research Ethics Review Board.

### Sample collection and processing

Blood (5 ml) was collected into gold top serum separator tubes (BD) by venipuncture and centrifuged at 1300 × g for 15 min to extract serum. Nasal flocked swabs were collected into universal transport media (COPAN Diagnostics). All samples were stored at -80C prior to analyses. Serum was inactivated by either heating at 56 °C for 30 min prior to ELISA, or by a combination of 56 °C and trypsin-heat-periodate treatment ^10^ prior to MNT and HAI assays.

### Microneutralization (MNT) assays

Patient sera were serially diluted 2-fold to create a 40 to 5120-fold dilution series using MEM supplemented with 1X Pen/Strep, 25 nM HEPES, and TPCK-treated trypsin (0.25 - 1 μg/mL) (Sigma-Aldrich). The cH5/1 N3 virus, engineered to express cH5/1, a fusion of the A/Vietnam/1203/04 H5 head domain with PR8 group 1 stalk and neuraminidase N3^9^, and propagated in 10-day-old embryonated chicken eggs, was incubated with dilutions of serum at a concentration of 400 PFU/well at RT for 30 min. The virus-serum mix was then added to MDCK cells plated in a 96-well plate at ∼90% confluency and incubated at 5% CO_2_ for 1 h at 37 °C. The plate was then washed and supplemented with DMEM containing 0.2 M L-glutamine, 100U/mL penicillin and 1μg/μL streptomycin overnight at 5% CO_2_, 37 °C, followed by fixation with 80 % acetone. Influenza virus nucleoprotein (NP) was measured using biotin-conjugated anti-NP antibody (EMB Millipore MAB8257B) and HRP-conjugated streptavidin (EMB Millipore). SigmaFast OPD (Sigma-Aldrich) was used as HRP substrate and luminescence was measured using the SpectraMax i3 plate reader at OD490. A MNT_50_ endpoint titer, serum dilution resulting in at least 50 % inhibition of infectivity, was defined as the reciprocal of the lowest serum dilution factor, for which normalized signal remained below the threshold of detection; endpoints outside the assay’s lower limit of detection (< 40) were assigned an endpoint titer of 20.

### Haemagglutination inhibition (HAI) assays

Serum was mixed with the A/California/04/2009 (Cal/09) H1N1 virus (a kind gift of Dr. Peter Palese, Icahn School of Medicine at Mount Sinai, NY) and incubated for 30 min at RT. Chicken erythrocytes were then added to the plate and incubated at 4 °C for 45 min. An HAI titer was defined as the highest serum dilution factor leading to HAI activity; samples without any detectable HAI activity were assigned a titer of “1”.

### ELISA assays

All viral antigens were made using the baculovirus expression system and constructs kindly gifted by Dr. Peter Palese and Dr. Florian Krammer (Icahn School of Medicine at Mount Sinai, NY). The cH6/1 HA construct was composed of a A/Mallard/Sweden/81/02 H6 head domain and a A/Puerto Rico/8/1934 (PR8) group 1 stalk ^9^. Briefly, construct sequences were cloned into pFastBac plasmids containing a C-terminal trimerization domain and hexahistidine tag, followed by transformation into DH10Bac bacteria. The resulting bacmids were transfected into Sf9 cells to generate viral stocks used to infect High Five insect cells. Viral proteins were then purified by Ni-resin affinity chromatography. 96-well plates were coated with 2 μg/mL of Cal/09 or cH6/1 HA, followed by 4 °C incubation overnight and blocking with 5% non-fat milk in PBS-T. Sera were added to the plate in a 2-fold serial dilution series ranging from 100-to 6400-fold. Mucosal samples were assayed undiluted. Plates were then incubated for 1 h at RT, HRP-conjugated goat anti-human IgG or IgA were added. SigmaFast OPD (Sigma-Aldrich) was used as HRP substrate and luminesence was measured using the SpectraMax i3 plate reader at OD490. A detection threshold was defined as 3 standard deviations above the average background signal. Serum IgA and IgG endpoint titers were defined as the highest serum dilution factor for which normalized OD490 values remained above the background threshold; endpoints outside the assay’s limit of detection (i.e. <100 or >6400) were assigned endpoint titer values of 50 and 25600, respectively. Mucosal ELISA readouts were reported as normalized OD490 values. Values <0 post-normalization were assigned a value of 0.

### Statistical analysis

Statistical analyses were performed using IBM SPSS v.27 and GraphPad Prism v.7. Two-sided Mann-Whitney U and Fisher’s exact tests were used to assess demographic differences among the participant groups. Data were scaled via log_2_ transformation. Geometric mean fold changes (GMFC) were calculated as ratios of geometric means (i.e. pre-/post-vaccination) for each parameter. Mucosal readouts equalling zero were substituted with assay-specific lowest limit of detection values for the GMFC and log_2_ calculations. Paired analyses were done using the paired t-test; differences between the LAIV and IIV groups were assessed using the unpaired t-test; correlation analyses were performed using the Pearson correlation coefficient test and lines of best fit were derived via linear regression.

## Data Availability

All data is provided in the manuscript

## ACKNOWLEDGEMENTS

The authors would like to thank the cRCT participants and study teams. This work was supported, in part, by a CIHR Project Grant (M.S.M.). M.S.M. was supported, in part, by a CIHR New Investigator Award and an Ontario Early Researcher Award. S.Y. was supported, in part, by a M.G. DeGroote Postdoctoral Fellowship.

## AUTHOR CONTRIBUTIONS

Conceptualization, M.S.M. and M.L. Investigation and formal analysis, M.S.M., S.Y., D.B.C., K.B.G., J.C.A., C.V., J.W., K.R., and V.T. Writing – original draft, M.S.M., S.Y., D.B.C. and K.B.G. Writing – review and editing, M.S.M., M.L., S.Y., D.B.C., K.B.G., J.C.A., C.V., J.W., K.R., V.T. Funding acquisition, M.S.M. and M.L.

